# Exploiting Molecular Basis of Age and Gender Differences in Outcomes of SARS-CoV-2 Infections

**DOI:** 10.1101/2021.05.23.21257669

**Authors:** Daniele Mercatelli, Elisabetta Pedace, Federico M. Giorgi, Pietro Hiram Guzzi

## Abstract

**Motivation:** Severe acute respiratory syndrome coronavirus 2 (SARS-CoV-2) infection (coronavirus disease, 2019; COVID-19) is associated with adverse outcomes in patients. It has been observed that lethality seems to be related to the age of patients. Moreover, it has been demonstrated that ageing causes some modifications at a molecular level.

**Objective:** The study aims to shed out light on a possible link between the increased COVID-19 lethality and the molecular changes that occur in elderly people.

**Methods:** We considered public datasets on ageing-related genes and their expression at tissue level. We selected interactors that are known to be related to ageing process. Then, we performed a network-based analysis to identify interactors significantly related to both SARS-CoV-2 and ageing. Finally, we investigated changes on the expression level of coding genes at tissue, gender and age level.

**Results:** We observed a significant intersection between some SARS-CoV-2 interactors and ageing-related genes suggesting that those genes are particularly affected by COVID-19 infection. Our analysis evidenced that virus infection particularly affects ageing molecular mechanisms centred around proteins EEF2, NPM1, HMGA1, HMGA2, APEX1, CHEK1, PRKDC, and GPX4. We found that HMGA1, and NPM1 have a different expression in lung of males, while HMGA1, APEX1, CHEK1, EEF2, and NPM1 present changes in expression in males due to aging effects.

**Conclusion:** Our study generated a mechanistic framework to explaining the correlation between COVID-19 incidence in elderly patients and molecular mechanisms of ageing. This will provide testable hypotheses for future investigation and pharmacological solutions tailored on specific age ranges.

## 1 Introduction

At the end of 2019 in Wuhan (China), medical facilities reported acute pneumonia cases with an unknown origin. Further analysis revealed that a novel coronavirus, named severe acute respiratory syndrome coronavirus 2 (SARS-CoV-2), was responsible for that disease, subsequently called coronavirus disease 2019 (COVID-19) [1, 2]. The clinical manifestations spanned from asymptomatic infection to severe pneumonia and a severe state of inflammation (molecularly characterised by a cytokine storm) leading to a fatal outcome [3, 4, 5, 6, 7, 8].

Starting from China, the virus spread in almost all other countries globally, causing infections and deaths. On 11th March 2020, the World Health Organisation (WHO) declared SARS-CoV-2 as a pandemic. Current data revealed that the impact of COVID-19 presents certain peculiar aspects in different nations that have been deeply investigated [9, 10]. Some authors hypothesised that virus mutations were responsible for these differences [11, 12, 13, 14]. Nevertheless, many independent studies agreed that the mutations might not have a primary role in explaining these differences [15, 16, 17].

Despite the lack of the individuation of the causes, there was a substantial agreement on the fact that the variation of the observed case fatality rate (CFR), i.e. the fraction of confirmed cases leading to fatal outcomes, ranging from 0 to 20% and beyond at country level, needs to be deeply investigated [18, 19, 20]. Among the other differences, we focused on observing that the infection is significantly more lethal in older people [21, 22, 23, 24, 25]. This consideration has also guided the optimisation of vaccination strategy [26].

Some studies have focused on the possible link between increased mortality rate and some characteristics of older people [27, 28]. In addition, these studies suggested the potential effect of the virus as a trigger activating the decompensation of other chronic conditions [29, 30, 31, 32]. Akbar et al., [33], discussed a possible link between the increased chronic inflammatory status occurring during ageing (termed “inflammaging” [34, 35]), and COVID-19 manifestation that causes the rise of inflammation.

Previous studies have also evidenced that the understanding of modification of molecular mechanism related to the ageing process (i.e. modification of gene expression, modulation of regulatory mechanisms) may reveal important insights about ageing [36]. Many studies contributed to identifying such ageing-related diseases despite the lack of having experimental data [37, 38, 39, 35, 40]. Computational predictions have also been made in [36, 41] giving both candidate genes and networks [42, 43].

Consequently, the study of the intersection between SARS-CoV-2 and ageing-related molecular alterations could augment the understanding of COVID-19, thus improving treatment options [44]. Bhattacharyya et al. presented a first analysis based on some preliminary public data reinforcing the rationale that such a possible link exists [45].

Six functional open reading frames (ORFs) in the SARS-CoV-2 genome encodes for the four main structural proteins, the Spike (S), Envelope (E), Membrane (M), and the Nucleocapsid (N), and ORF1a/ORF1b, which contain information for the replicase–transcriptase complex formed by 16 non-structural proteins (NSP1–NSP16). The SARS-CoV-2 genome also contains 9 accessory factors from sub-genomic ORFs (Orf3a, 3b, 6, 7a, 7b, 8, 9b, 9c and 10) [46]. We investigated the relationships and interactions between these viral components and age-related factors and observed a significant overlap between SARS-CoV-2 and ageing group genes’ interactors. Furthermore, we looked at a network-level scenario [43], by considering possible regulatory mechanisms that may be altered [47, 48]. These observations support previous reports that SARS-CoV-2 also involves a vascular and multiorgan failure in severe COVID-19 [49].

Starting from these considerations, we hypothesised that SARS-CoV-2 interacting proteins (and genes) might show an overlap with human ageing-related genes higher than chance. Therefore, the infection may deregulate these mechanisms that can be already impaired in older adults, causing severe outcomes. We downloaded public available interaction data from Guzzi et al. [50] and Gordon et al., [51]. Then we considered the interacting partners that were annotated as *ageing* genes in MSigDB [52] ad we also considered the expression at tissue and sex levels extracting data from GTeX database [53]. We verified a significant fraction of interacting partners of SARS-CoV-2 involved in ageing. These genes are also expressed in lung and the expression is modulated by age and sex. We also observed that these genes are expressed in adipose tissue (as reported in Supplementary Material). The workflow of the experiment is depicted in Figure 1.

**Figure 1:**
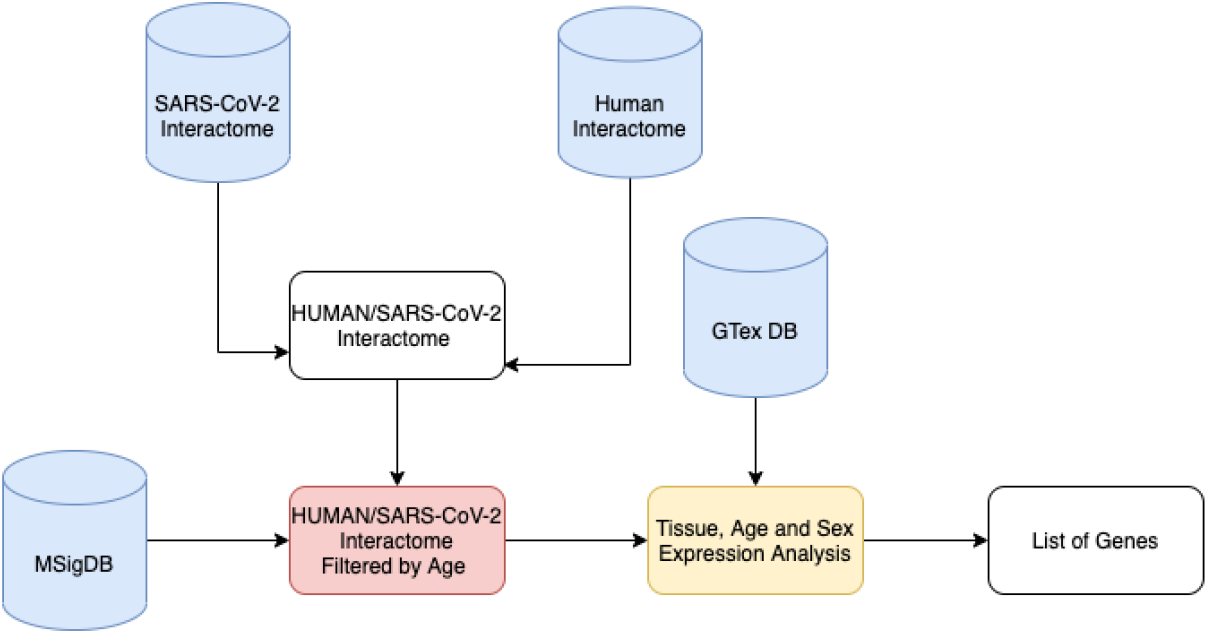
Workflow of the experiment. We downloaded public available interaction data from previous studies. We built the integrated human/SARS-CoV-2 interactome. In parallel we downloaded the list of genes annotated with *ageing* keywords as in MSigDB database. Then, for each SARS-CoV-2 protein we calculated the probability that it contains human interactors annotated with with *ageing* keyword. We obtained a list of SARS-CoV-2 proteins containing a significant number of interactors related to aging. Then we calculated the intersection of these sets (*core interactors*) obtaining a list of eight human proteins. For each core interactor, we also considered the expression at tissue level extracting data from GTeX database. We verified that there exist a significant fraction of interacting partners of SARS-CoV-2 that are involved in ageing and that are particularly expressed in lung and in adipose tissue.

## 2 Methods

### SARS-CoV-2 Interaction Map

We considered the SARS-CoV-2 protein interaction map provided by Gordon et al., [51], and by Guzzi et al., [50]. Both works provided data about 26 of the 29 SARS-CoV-2 proteins behaviour in human cells by identifying the human proteins that physically associated with each of the SARS-CoV-2 proteins using affinity-purification mass spectrometry. They found high-confidence protein-protein interactions between SARS-CoV-2 and human proteins; they also provided data about possible interactions with an associated reliability score. We considered both high and low confidence interactions.

### Databases

We first defined and labelled genes related to the ageing process as *ageing*. Then, we considered data provided from the GTEx dataset containing genes positively and negatively correlated with human age [53]. We gathered data from the GenAge dataset that derived human genes by projecting sequence orthologs in model organisms. We also considered the MSigDB gene set collections, which summarized gene information associated with *ageing* collected from 70 different studies. We selected datasets reporting experiments from homo sapiens since orthologs’ projection may produce not reliable results for ageing as described in [36].

We used the Search Tool for the Retrieval of Interacting Genes/Proteins database (STRING) [54] that is a freely available repository storing both physical and functional association among proteins. Users may search the database through a web interface by specifying a protein identifier or inserting the primary sequence. We queried the database using the identifiers of the nodes of each subnetwork. We used medium confidence as the minimum confidence score for each interaction and *all* for the sources of interactions. We searched the GTeX Portal [55] using the previously described list of gens. We obtained the expression of those genes in a heat map that shows expression across all GTEx tissues. Gene Ontology analysis was performed by using Gene Ontology web portal [56] while Reactome Database was used for identifying related pathways [57].

### Bioinformatic Analysis

We selected all known SARS-CoV-2 interacting partners. Then, we measured the intersection between this list of interactors and the ageing-related genes for each viral protein and estimated the probability that this intersection is higher than chance by Fisher’s exact test.

We also tested the significance of the difference in the expression of EEF2, NPM1, HMGA1, HMGA2, APEX1, CHEK1, PRKDC, and GPX4 due to age (we considered six different classes), sex, and tissue. We used a Wilcoxon Test for testing difference in the expression among classes (since the expression of genes is not gaussian as reported by a Shapiro test). In addition, the difference among age classes is evaluated using a Kruskal Wallis test.

## 3 Results

### 3.1 Network Analysis

For each viral protein we select the human interactors. The analysis revealed that only ten viral proteins (M, NSP2, NSP4, NSP6, NSP11, NSP13, Orf3a, Orf7a, Orf8, and Orf9c) have interactors with a significant overlap with respect to ageing related proteins, as summarised in Table 1. Then, we considered those that are enriched for ageing in a significant way. Finally, we intersected all these sets and we obtain a core set of eight proteins: EEF2, NPM1, HMGA1, HMGA2, APEX1, CHEK1, PRKDC and GPX2 (*core interactors* hereafter) as reported in Figure 2 (see supplementary for the list of interactors for each viral protein, integrated with the topological characteristics of the induced subnetwork in the human interactome).

**Table 1:** P-Values of the enrichment. For each protein we report the significance of the enrichment. A p-value lower than 0.01 means that the interactors are significantly related to ageing. (NS stands for not significant)

| Viral Protein | P-Value | Viral Protein | P-Value |
| --- | --- | --- | --- |
| Spike | NS | E | NS |
| **M** | **6.84E-02** | N | NS |
| NSP1 | NS | **NSP2** | **0.00182** |
| NSP3 | NS | **NSP4** | **8.32E-03** |
| NSP5 | NS | **NSP6** | **0.002676** |
| NSP7 | NS | NSP8 | **0.003421** |
| NSP9 | NS | NSP10 | NS |
| **NSP11** | **0.0001862** | NSP12 | NS |
| **NSP13** | **0.002573** | NSP14 | NS |
| NSP15 | NS | NSP16 | NS |
| **Orf3a** | **5.06E-03** | Orf3b | NS |
| Orf6 | NS | **Orf7a** | **0.0001791** |
| Orf7b | NS | **Orf8** | **0.0006913** |
| Orf9b | NS | **Orf9c** | **1.50E-02** |
| Orf10 | NS |  |  |

**Figure 2:**
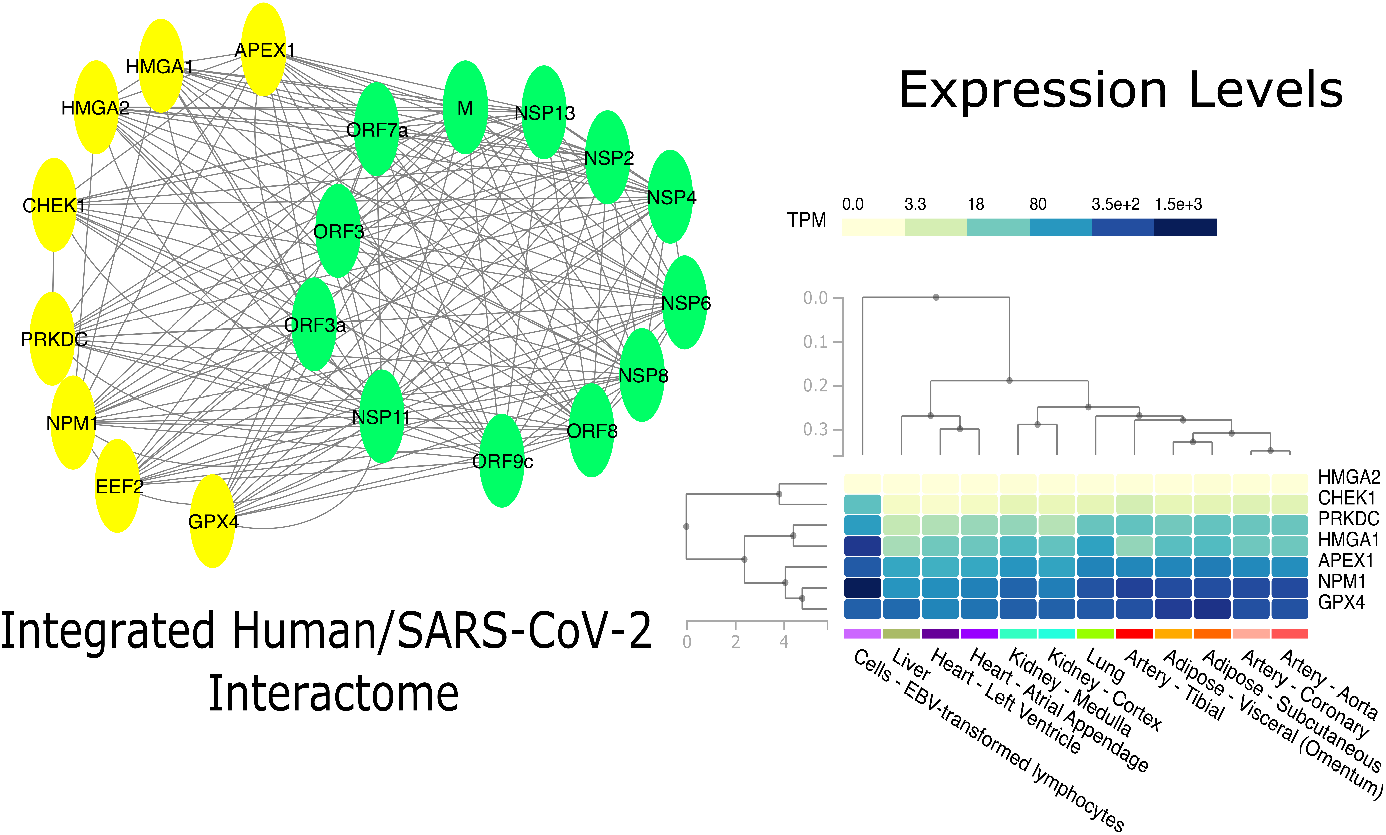
Figure shows tissue level analysis of this work. The Network analysis contributed to find a set of human proteins (yellow nodes) related to aging that interact with many SARS-CoV-2 proteins (green nodes). The analysis of the expression of the related genes at tissue level revealed that all these genes are expressed in lung, as well as in other human tissues. Expression level are presented as TPMS.

The Gene Ontology analysis reveals that the whole network is enriched with the following terms: (GO:0090402) oncogene-induced cell senescence, (GO:0035986) senescence-associated heterochromatin focus assembly, (GO:2000774) positive regulation of cellular senescence, (GO:2000773) negative regulation of cellular senescence, (GO:2000772) regulation of cellular senescence. The analysis of Reactome DB reveals that the subnetwork is associated with following pathways: Formation of Senescence Associated Heterochromatins Foci (HSA2559584), Host interactions of HIV factors.

### 3.2 Expression Analysis

We searched the GTeX database for the expression (of *core interactors* as reported in Figure 2 expressed as TPM (Transcripts Per Million).

We found that all the interactors are expressed in lung as well as in other human tissues (supplementary materials contain the expression levels of these genes in different human tissues). In order to assess the different outcomes between male and female we focused on lung tissue and we compared the expression of these core interactors in male and female as reported in 3. Since data are not normally distributed, (as given by Shapiro Test), we measured the difference of expression in male/female class. We evidenced a significant difference tested by using a Wilcoxon Test for NPM1 and HMGA1 genes that are significantly downregulated in males, without considering age as reported in Figure [**?**].

We also explored the trend of the core interactors focusing on lung tissue and six different classes of age (20-29, 30-39, 40-49, 50-59, 60-69, 70-79). We found that there is a significant difference considering age groups for HMGA1, APEX, CHEK1, EEF2, and NPM1 (p ≤ 0.05 as evidenced by a Kruskal Wallis test). Figure 4 report this trend.

**Figure 3:**
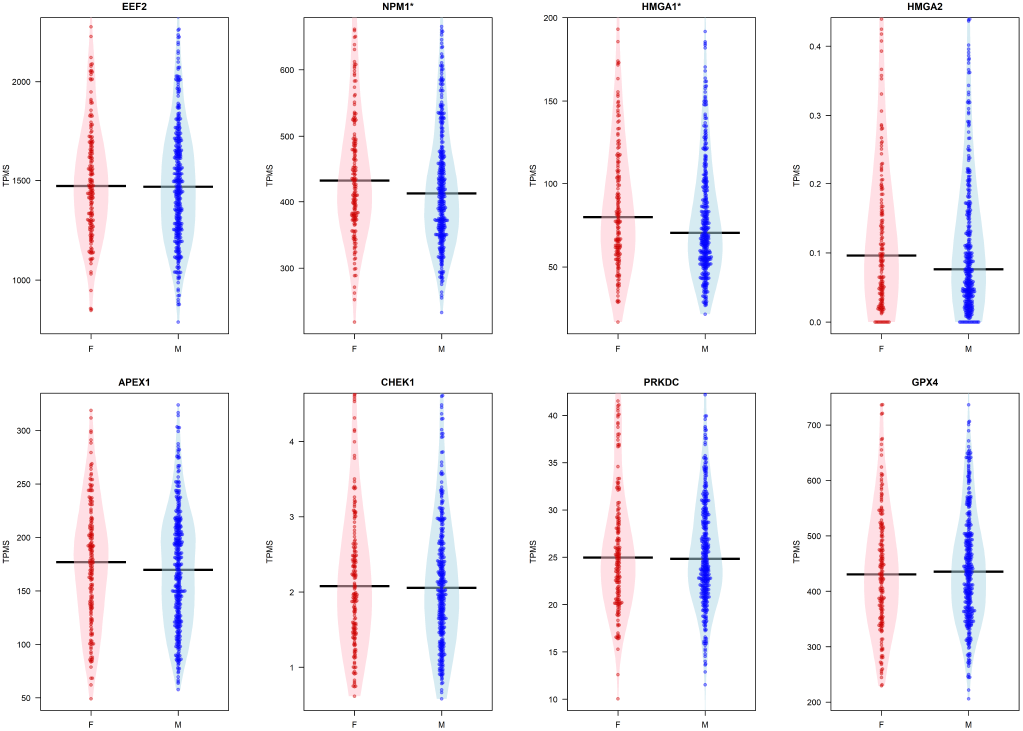
Figure reports box plot of the expression of the eight core genes grouped by sex in the lung tissue. The evidences a significant difference tested by using a Wilcoxon Test for NPM1 and HMGA1 genes.

**Figure 4:**
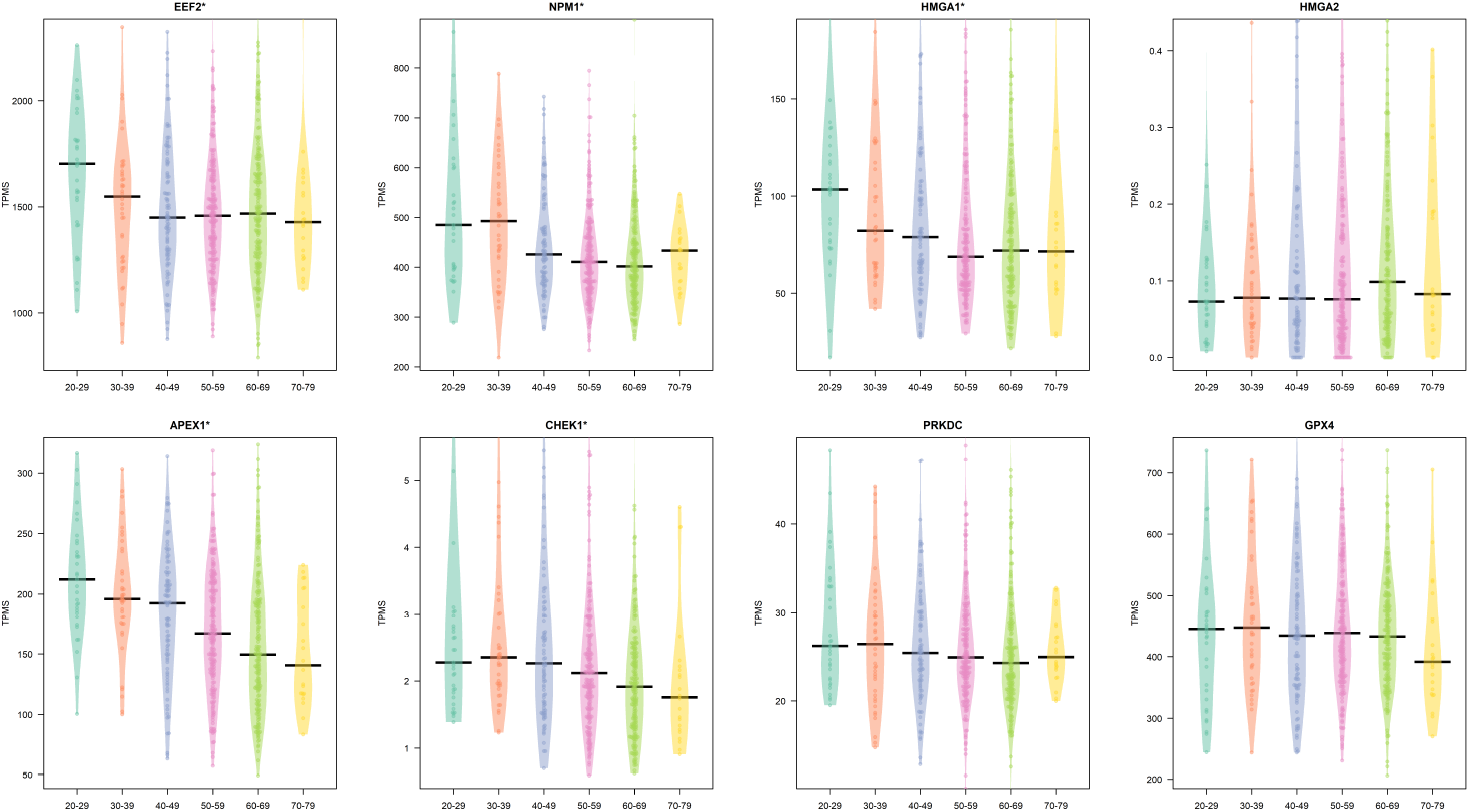
Figure reports the difference of the expression of the core genes in lung tissue in different age classes. A * on top of the plot means a significant difference (p ≤ 0.05 as evidenced by a Kruskal Wallis test)

We then subdivided these groups considering sex. We found that that females present only the age dependent modulation of APEX1, while in males all the previous genes are modulated considering age groups as reported in Figures 6 (females) and 5 (males) (p ≤ 0.05 as evidenced by a Kruskal Wallis test).

## 4 Discussion

As introduced before, deaths from COVID-19 occur predominantly among older adults. COVID-19 also appears to be more lethal for men rather than women. This characteristic has been found in China, as well as in Europe and in the United States of America, [58].

Starting from this observation, we tried to explain the molecular basis of this phenomenon. Next, we recall that ageing is a heterogeneous process that presents differences among individuals. In particular, age-related changes impact many organs producing possible multi-organ failures even showing many inter-individual differences. Beyond these differences, we tried to explain how the age-related changes at the molecular level can be relevant to COVID-19 pathology.

To achieve this goal, we integrated interactomics and expression data related to COVID-19, age and sex. We started from SARS-CoV-2 interactors, and we isolated age-related from those. Then we considered the expression value of these genes, and we further investigated the trend of changes of these genes in age and sex groups. In summary, we identified a set of statistically significant interactors for the aging process: EEF2, NPM1, HMGA1, HMGA2, APEX1, CHEK1, PRKDC, and GPX4. As reported in Figure 8 we found some interesting changes of these genes considering tissue, age and sex groups. We also found that NPM1 and HMG1 are downregulated in males (statistically significant regulation); while HMGA2 is slightly downregulated in males (not significantly) (Figure 3).

We also found some statistically relevant changes in age for EEF, NPM1, HMGA1, APEX1, and CHEK1 for males (Figure 5), and for APEX1 in Females (Figure 6).

**Figure 5:**
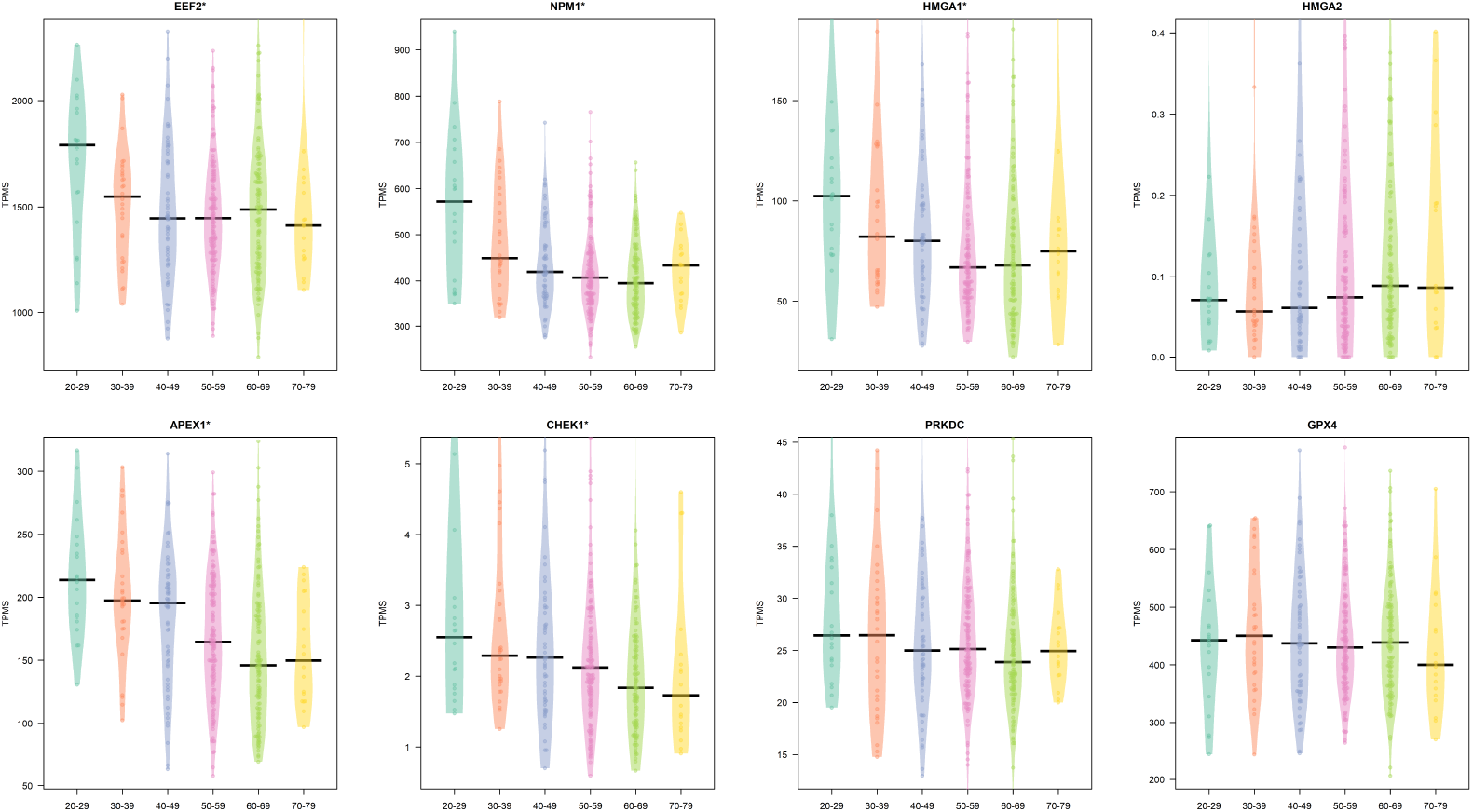
Difference in the expression in lung tissue by age classes in males. Expression is reported as TPM.A * on top reveals a modulation in groups.

**Figure 6:**
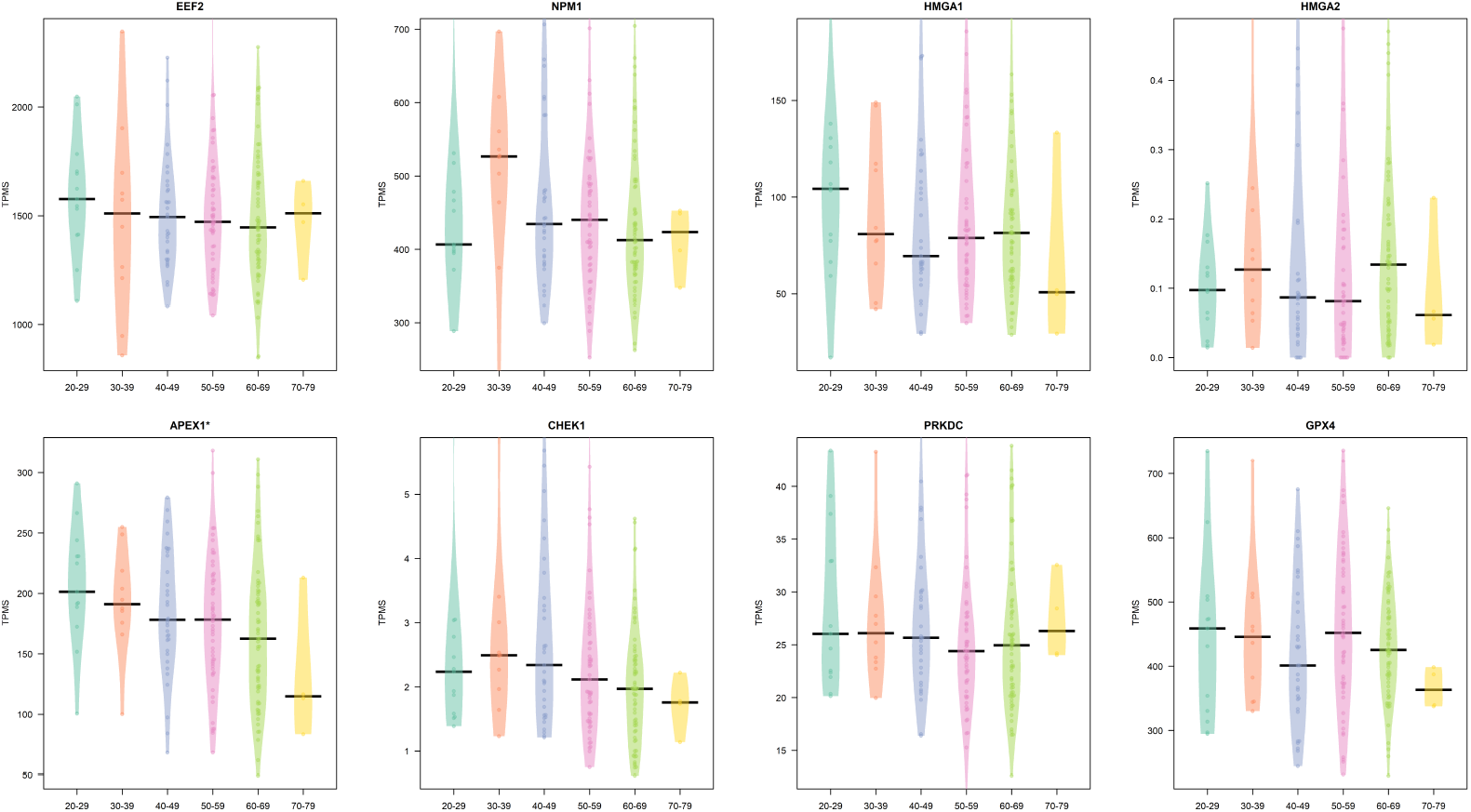
Difference in the expression in lung tissue by age classes in females. Expression is reported as TPM. A * on top reveals a modulation in groups.

**Figure 7:** Expression of core interactor in males and females grouped by age class.

Our findings are coherent with the observation and the literature related to COVID-19.

As investigated in [59] ageing is characterised by the decline of the immune function. Older adults are not immuno-deficient, but often the immune system’s response is not sufficient to be effective again antigens. This effect is particularly evident when they are subject to novel antigens. For example, it is known that both responses to influenza and vaccination are not efficient in the elderly [60, 61]. Moreover, the elderly accumulate inflammatory mediators in tissues (inflammageing process), that may occur by the accumulation of DNA lesions that, in turn, triggers the increased production of inflammatory mediators [62]. In parallel, the link between COVID-19 and the suppression of the immune system has been observed in [63]. Authors found that many proteins related to the immune response were modulated, causing the possible suppression of such a system.

HMGA1 and HMGA2 genes encode four proteins (HMGA1a, HMGA1b, HMGA1c, and HMGA2) belonging to the High-mobility group A (HMGA) protein family [64]. All the proteins bind AT-rich regions in DNA and modulate gene expression by acting as transcription factors. Literature reports that HMGA1 has critical roles in tumorigenesis and the progression of various cancers. However, the role of HMGA1 in COVID-19 has not explored in the past. We now demonstrate that HMGA1 is a SARS-CoV-2 interactor. It is differently regulated in males and the elderly, so these differences in expression are associated with poor prognosis in the found classes (elderly and males). It has been shown that HMGA1 induces inflammatory pathways early in many cancers and pathways involved in stem cells, cell cycle progression, and its dysregulation causes aberration in pathways of stem cells, cellular development, and hematopoiesis [65]. Our results provide insight into HMGA1 function during COVID-19 pathogenesis that could serve as therapeutic targets in human people with aberrant HMGA1 expression.

Similarly to HMGA1, the Nucleophosmin (NPM1) is also downregulated in males. NPM1 is related to DNA and cell cycle control such as ribosome biogenesis, protein chaperoning, centrosome duplication, histone assembly, and cell proliferation [66, 67]. Previous studies investigated the age incidence of acute myeloid leukaemia with mutated nucleophosmin (NPM1) [68, 69], while there are no studies related to these mutations and other diseases. In [**?**] the impact of NPM1 modification in older patients has been investigated for AML, suggesting a worse prognosis for older patients due to NPM1 changes. Therefore, this work may stimulate further studies in such a direction. The interaction between NPM1 and the nucleocapsid protein of the previous SARS-CoV is known to affect the viral particle assembly [70, 71, 72]. The role of NPM1 and of Histone H2AX targeted by other viral proteins has also been reported in other viruses such as Epstein-Barr and KSHV as a common strategy to manipulate translation and to promote virus latency [73, 74].

Moreover, for older men, the scenario is complicated by the down-regulation of EEF2, APEX1 and CHEK1.

The dysregulation of EEF2 may cause the accumulation of DNA damage or, on the protein levels accumulating errors in the down-stream cascade of mechanisms [75]. The role of EEF2 in severe cases of COVID-19 has also been elucidated in [63]. Thus our study provides another evidence. Moreover, this protein is targeted together with the Eukaryotic translation initiation factor 2 subunit 1 (EIF2S1) by Orf3a, Orf8, NSP2, NSP6, NSP11, NSP13, indicating a possible role of the virus to promote viral translation over cellular translation [76]. In [77] the synergistic downregulation of both APEX1 and NPM1 has been clearly observed in oligodendrocyte cells in relation to ageing. (APEX1) plays an essential role in the cellular response to oxidative stress. APEX1 has a major role in DNA repair and in redox regulation of transcription factors [69]. CHEK1 is targeted together with CDK1 by many SARS-CoV-2 interactors (NSP2, NSP4, NSP11, NSP13) and with CDKN2A (Orf3, NSP13), suggesting an additive effect on the disruption of pathways of apoptosis mediated by TP53 [78] yet dis-regulated by both senescence and ageing. [79].

Differently, for females, we found only the age-dependent modulation of APEX1, suggesting that females may have less factor risk than males. This is coherent with the observation that COVID-19 has a lower CFR rate in females.

In parallel, in supplementary material, we report that that core interactors are also significantly overexpressed in adipose tissue, therefore suggesting a second factor of co-morbidity. Changes in adipose tissue promote a chronic state of low-grade systemic inflammation on a phenotypic level, thus increasing the risk of age-associated diseases [35, 80]. We here report that core interactors are expressed in adipose tissue, suggesting a possible role that should be further investigated.

## 5 Conclusion

We applied a bioinformatic analysis to perform a qualitative analysis of mechanisms of infection by SARS-CoV-2 in elderly people.

Several studies have shown in the past the modifications of genes and proteins that occur in elderly people. Other studies have partially elucidated the mechanism of infections and the dysregulated pathways in COVID-19 patients.

We detected a statistically significant overlap between SARS-CoV-2 interacting proteins and those related to ageing, suggesting a potentially different response in elderly people. Our analysis evidenced that virus infection particularly affects ageing molecular mechanisms centred around proteins EEF2, NPM1, HMGA1, HMGA2, APEX1, CHEK1, PRKDC, and GPX4. We also found that these genes are expressed in lung. Finally we found that there is a significant difference in the expression considering both age and sex. These results will provide an important molecular basis for understanding the mechanism of infections and will shed light on infection progression. The limitation of this study is that the dataset is correlative, and thus it should be confirmed by *in vivo* experiments.

**Figure 8:**
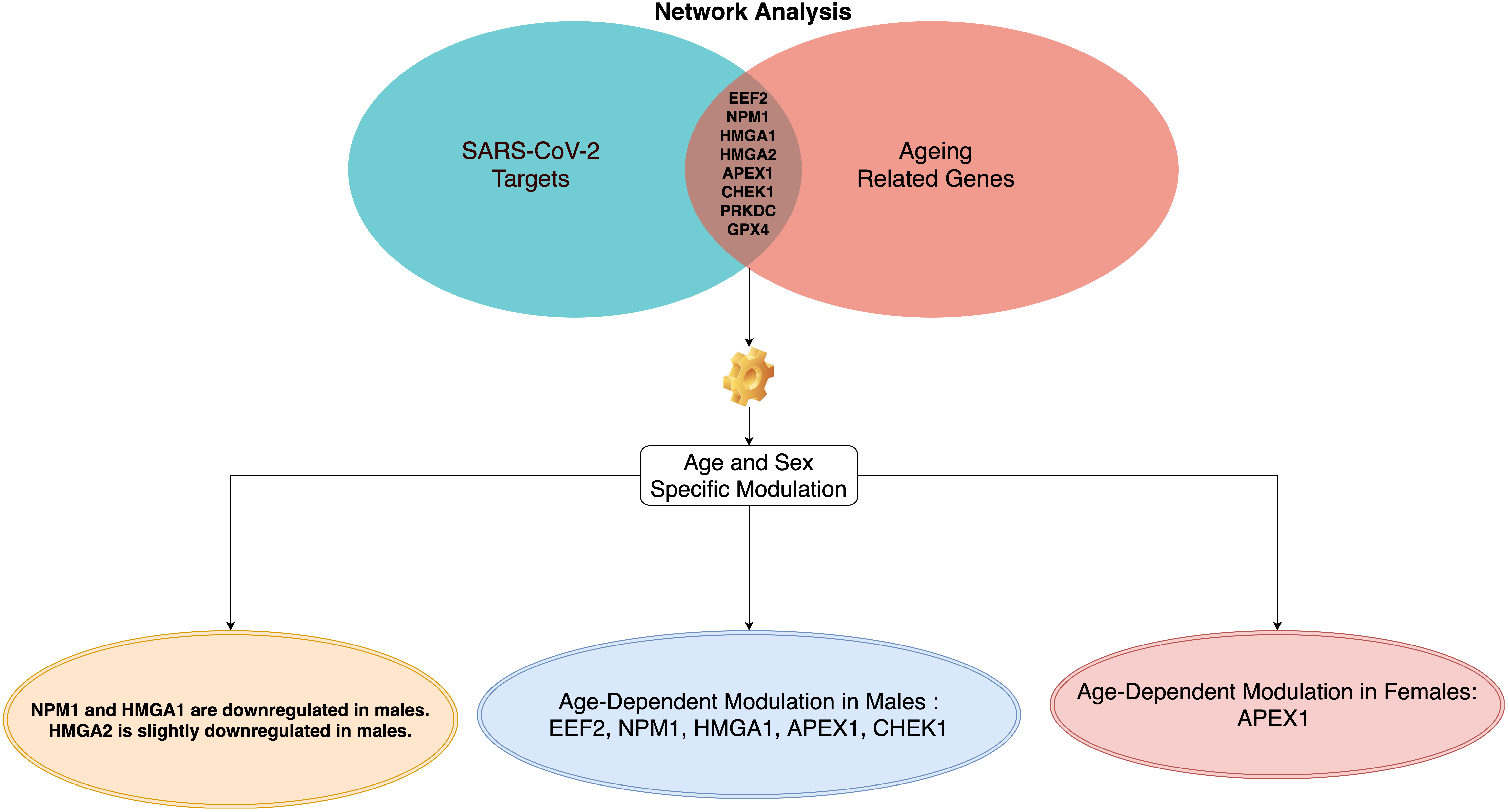
Figure summarises main results of the work. Network analysis found that there exists eight proteins related to ageing that are also all targeted by ten SARS-CoV-2 proteins. The analysis of the expression of their genes revealed that there exist difference on the expression of these genes considering both age and sex.

## Data Availability

All data used have been downloaded from public databases we cited in the manuscript.

## Appendix

### Conflict of Interests

Authors state that they do not have conflict of interest.

### Author Contribution

F.M.G and P.H.G. conceived the main idea of this manuscript. D.M. performed the experimental analysis. Both participated to the experimental phase and on the discussion of the results. E.P. participated to writing of Discussion Section and also validated the clinical aspects of this work. All authors read and approved the manuscript.

## Acknowledgement

Author thank C.V. Cannistraci for useful suggestions and discussion during the preparation of this work.

## 6 Key Points

- A network-based analysis identified some molecular mechanisms that could play a role in the SARS-CoV-2 molecular aetiology and ultimately affect COVID-19 outcome.
- Our analysis evidenced that virus infection particularly affects ageing molecular mechanisms centred around proteins EEF2, NPM1, HMGA1, HMGA2, APEX1, CHEK1, PRKDC, and GPX4.
- We found an age dependent modulation of EEF2, NPM1, HMGA1, APEX1 and CHEK1 in lung tissue of males.
- We found a age dependent modulation of APEX1 in females.
- Our study generated a mechanistic framework aiming at explaining the correlation between COVID-19 incidence in elderly patients and molecular mechanisms of ageing considering differences by age and sex.

## 7 Supplementary Materials

File Supplementary.tex

